# Knowledge and representations of the placenta in a French population of women *post-partum*

**DOI:** 10.1101/2024.10.11.24315298

**Authors:** Sébastien Riquet, Jade Busca, Carole Zakarian, Pierre Le Coz, Jean-Louis Mege, Soraya Mezouar

## Abstract

In many cultures, the placenta is considered to be the twin brother, counselor or guardian angel of the baby throughout pregnancy and is the subject of many beliefs and practices. In France, it is better known from a biomedical point of view including the protocols related to its disposal as biomedical waste in the hospital environment. Our study aims to evaluate the knowledge and representations of the placenta before being discarded by health personnel who assisted the parturient at birth. Our results show a lack of knowledge and misconceptions about the placenta and its different functions. Nevertheless, mothers consider it essential to their children’s development and are ready to give it to science "if it can help" once the birth is complete. Information and documentation on the placenta among pregnant women are necessary for better management of childbirth and practices, especially for mothers who wish to give birth at home or in a delivery center.

## Introduction

The placenta is essential for gestation and physiological or surgical birth as summarized by this statement "*no placenta no baby and no baby no placenta*" [1]. During the intrauterine life of the fetus, it plays a critical role in transmitting the oxygen, ensuring the energy supply and allowing the elimination of its waste [2]. In many cultures, the placenta is considered throughout pregnancy as the baby’s twin brother, counselor, or guardian angel and is the subject of many beliefs and practices [3,4]. In France, it is better known from the biomedical point of view including the protocols related to its disposal as a biomedical waste in the hospital environment [5]. There are concerns in some French overseas territories (Polynesia, Mayotte) and in certain communities established in France sometimes regarding the transmission of practices linked to beliefs around the placenta [6,7]. At the time of delivery, the term “delivery” marks the end of the life of this transitory organ. At the time of delivery, the placenta is removed from the eyes of parents in maternity wards because delivery is “repulsive” to modern man [8].

In France, the value of the placenta is only informative: knowledge of its structure, functions and pathologies that occur during pregnancy. The placenta and the fetal appendages (umbilical cord, amniotic membranes) are the body of important biomedical knowledge, and many specific scientific publications are dedicated to it. Since the appearance of banks to store stem cells and umbilical cord blood cells, the symbolic value of the placenta is now seen through prism of genetic technology by researchers and families who consider them for their therapeutic properties. In several other cultures, the placenta is first the object of various rites that apply after birth (burial, drying, etc.) [9]. However, the placenta remains poorly known and even ignored by most pregnant women because it is complex and difficult to access in terms of its origin, its constitution. Parents also do not know that the placenta is an organ of the same karyotype as the fetus [10].

The knowledge and representations of women who gave birth have been collected on this organ which allows the implantation of the embryo, the course of the pregnancy and participates in the induction of delivery before being thrown away by health staff members who assisted the parturient at the birth. However, it is precisely because the placenta is discarded as a waste that we wanted to explore the knowledge of this organ with mothers who have just given birth and assess their interest in donating it for research.

## Materials and Methods

A non-interventional phenomenological study was carried out between September 1^st^ 2019, and May 31^th^ 2020 at the “Assitance Publique-Hopitaux de Marseille” (APHM, Nord and Conception hospitals) and the maternity ward of the Beauregard clinic in Marseille. The state of health emergency (from March 23^th^ to July 10^th^, 2020) decreed a generalized confinement of the French population (from March 17^th^ to May^th^ 11) to face the Coronavirus disease 2019 (Covid-19) pandemic, an appointment was made with the mothers, before their early discharge from the maternity ward, to carry out the telephone interview with them according to their availability at home. Semi-structured interviews were conducted with women who had given birth, in the first week of postpartum, following three stages: 1) a survey of the sociologic demographic and economic characteristics of the participants by the interviewer; 2) the achievement of drawing production by the mother; 3) an interview with the mother following an interview grid.

On the one hand, the data collection tools were composed of graphic production to solicit "brain mechanisms". At the beginning of the interview, before starting the questionnaire, a graphic production was requested from mothers to evaluate their specific cognitions on the placenta and fetal appendages. It is recognized that humans have an innate ability to graphically represent elements that they understand [11]. The presence of the placenta was assessed first. Next, the drawing productions were scored on five points. The presence of each of the following elements was assigned a point: the uterus, the placenta, the umbilical cord, the amniotic membranes and the amniotic fluid.

On another hand, a semi-structured interview permitted to collect oral knowledge and representations that the mothers have about the placenta during pregnancy and after delivery. Their interest and expectations of the placenta were also discussed during the interviews [12].

The inclusion criteria were: primiparous or multiparous pregnancies that lead to birth by vaginal or by caesarean section. Age, nationality, origin, religion, level of education, professional category, marital status, participation in “Birth and Parenthood Preparation” (BPP) courses, parity, mode of delivery and finally the presence of complications during pregnancy and/or childbirth were taken account. Minor mothers and protected adults, those who did not have a sufficient understanding of the French language to participate in an oral interview or who did not wish to participate in the study were not included.

Inclusion kits consisting of a study information leaflet, consent forms in two copies, a questionnaire and the interview grid, blank sheets of paper and a pen to make a drawing were put together to conduct the study. When the interviews were conducted by telephone, the mother produced her drawing at home and then sent a photo of her drawing by message to the interviewer’s mobile phone. The interviews lasted an average of 40 minutes. They were recorded and transcribed verbatim by one of the investigators, then the recordings were deleted.

The qualitative content analysis, by theme, of the data collected by the interview was done according to the Bardin method [13]. Initially, the interviews were subjected to an initial codification by two independent researchers. Then, in a second step, a confrontation of these codifications was practiced jointly by the researchers. Each of the categories and subcategories was validated following decision-making after exchanges and discussions between the researchers.

This study was approved by the Ethics Committee of Aix-Marseille University (n°2021-09-07-14).

## Results

### Study population

Sixty drawings and interviews, coded from E1 to E60, were carried out. Two-thirds of the interviews were conducted face-to-face between the mothers and the interviewer and for a third remotely by telephone interview during the pandemic period. The face-to-face interviews took place in the postpartum departments of the maternity in Marseille.

The sample of the study (n=60) was composed of 37 primiparous mothers (62%) and 23 multiparous (38%). Sixty percent of the mothers were French and 40% of foreign origin including Algeria (12%), Comoros (8%) and Armenia (7%). Fifty two percent were atheists, and the believers were divided between: Christian (32%) and Islam (17%). Their level of education was mostly higher than the high school diploma (67%) and the most represented professional categories were those of employees or managers (37%) and those with a health profession (22%). Most of them lived with the father (or second parent) of the child (88%), who were married or cohabiting. BPP sessions were attended by 60%, of whom 78% were primiparous and only 30% by multiparous women. They were largely non-smokers (85%), they did not have any complications during pregnancy (63%); three had a complication related to the placenta (low incineration, T1 detachment, accreta), they gave birth normally vaginally (70%) and five had an artificial delivery-uterine revision (AD-UR) (**Table 1**).

**Table 1.** Characteristics of mothers (n=60), without (n=46) and with (n=14) the drawn placenta. Birth and Parenthood Preparation (BPP) courses.

| **Characteristics n (%)** | **Total population**<br>**60 (100)** | **Without placenta**<br>**46 (76,66)** | **With Placenta**<br>**14 (23,33)** |
| --- | --- | --- | --- |
| **Age (year)** |  |  |  |
| 18-35 | 46 (76,67) | 33 (71,73) | 12 (85,71) |
| > 35 | 14 (23,33) | 13 (28,26) | 2 (14,28) |
| **Parity** |  |  |  |
| Primiparous | 37 (61,67) | 28 (60,69) | 9 (64,28) |
| Multiparous | 23 (38,33) | 18 (39,13) | 5 (35,71) |
| **Origin** |  |  |  |
| France | 36 (60,00) | 27 (58,69) | 9 (64,28) |
| Comoros/Mayotte | 5 (8,33) | 5 (10,86) |  |
| Bulgaria | 1 (1,67) | 1 (2,17) |  |
| Spain | 2 (3,33) | 2 (4,34) |  |
| Armenia | 4 (6,67) | 3 (6,52) | 1 (7,14) |
| Egypt | 1 (1,67) | 1 (2,17) |  |
| Algeria | 7 (11,67) | 4 (8,69) | 3 (21,42) |
| Italy | 1 (1,67) | 1 (2,17) |  |
| Senegal | 1 (1,67) |  | 1 (2,17) |
| Turkey | 1 (1,67) | 1 (2,17) |  |
| Kenya | 1 (1,67) | 1 (2,17) |  |
| **Religion** |  |  |  |
| Atheist | 31 (51,67) | 20 (43,47) | 11 (78,57) |
| Christian | 19 (31,67) | 17 (36,95) | 2 (14,28) |
| Islam | 10 (16,67) | 9 (19,56) | 1 (7,14) |
| **Level of education** |  |  |  |
| < High school diploma | 11 (18,33) | 8 (17,39) | 2 (14,28) |
| High school diploma | 9 (15,00) | 8 (17,39) | 1 (7,14) |
| Higher education | 40 (66,67) | 30 (65,21) | 11 (78,57) |
| **Professional Category** |  |  |  |
| Healthcare Professional | 13 (21,67) | 8 (17,39) | 4 (28,57) |
| Employee or manager | 22 (36,67) | 18 (39,13) | 4 (28,57) |
| Senior manager - liberal profession | 4 (6,67) | 4 (8,69) |  |
| Unskilled worker or employee | 15 (25,00) | 12 (26,08) | 3 (21,42) |
| Unemployed | 6 (10,00) | 4 (8,69) | 3 (21,42) |
| **Situation conjugale** |  |  |  |
| Alone | 7 (11,67) | 7 (15,21) |  |
| In a couple | 24 (40,00) | 19 (41,30) | 5 (35,71) |
| Married | 29 (48,33) | 20 (43,47) | 9 (64,28) |
| **Participation in BPP sessions** |  |  |  |
| Yes | 36 (60,00) | 27 (58,69) | 9 (64,28) |
| No | 24 (40,00) | 19 (41,30) | 5 (35,71) |
| **Smoker** |  |  |  |
| Yes | 9 (15,00) | 8 (17,39) |  |
| No | 51 (85,00) | 38 (82,60) | 14 (100) |
| **Complications during pregnancy** |  |  |  |
| Yes | 25 (41,67) | 15 (32,60) | 7 (50) |
| No | 35 (58,33) | 31 (67,39) | 7 (50) |
| **Mode of delivery** |  |  |  |
| Vaginal delivery | 42 (70,00) | 33 (71,73) | 11 (78,57) |
| Caesarean section | 18 (30,00) | 13 (28,26) | 3 (21,42) |
| **Artificial delivery-Uterine revision** |  |  |  |
| Yes | 5 (8,33) | 5 (10,86) |  |
| No | 55 (91,66) | 41 (89,13) | 14 (100) |
| **Drawing score** |  |  |  |
| 0/5 | 17 (28,33) | 17 (36,95) |  |
| 1/5 | 17 (28,33) | 17 (36,95) |  |
| 2/5 | 14 (23,33) | 7 (15,21) | 7 (50) |
| 3/5 | 9 (15) | 5 (10,86) | 4 (28,57) |
| 4/5 | 2 (3,33) |  | 2 (14,28) |
| 5/5 | 1 (1,67) |  | 1 (7,14) |

### Drawings

Seventy seven percent of mothers did not draw the placenta. When it was drawn, it was almost always associated with the umbilical cord and well connected to the umbilicus of the fetus. Most mothers had a drawing-average graphic score (80%). Twenty eight percent had the lowest drawing scores between zero and one. Only one reached the maximum of five points (**Table 1** and **Table 2**). The uterus was the most represented element and most often associated with the umbilical cord. Two mothers did not produce a fetus. When shown, it was positioned in a podal position. The cephalic position came in second place (37%) ahead of the transverse position (13%) (**Table 2**).

**Table 2.** Graphic scores and details of the productions (the number of times the element or combination of elements was found is given in parentheses) including (**A**) entire drawing production, (**B**) fetal position and (**C**) Umbilical cord implantation. AF: amniotic fluid, AS: Amniotic sac.

**A.**
| Score (N) | Drawing production |
| --- | --- |
| 0/5 (17) |  |
| 1/5 (17) | Uterus (11)<br>Cord (5)<br>AF |
| 2/5 (14) | Uterus + cord (6)<br>Uterus + placenta<br>Uterus + AF<br>Placenta + cord (6) |
| 3/5 (9) | Uterus + cord + placenta (4)<br>Uterus + cord + AS (3)<br>Uterus + cord + AF<br>Uterus + AF + AS |
| 4/5 (2) | Uterus + placenta + cord + AS (2) |
| 5/5 (1) | Uterus + placenta + cord + AS + AF |

**B.**
| N | Fetal position |
| --- | --- |
| 28 | Caudal |
| 22 | Cephalic |
| 8 | Transverse |
| 2 | Absent fetus |

**C.**
| N | Umbilical cord implantation |
| --- | --- |
| 11 | Placenta – foetal ombilic |
| 4 | Uterus – foetus |
| 2 | Uterus – foetal ombilic |
| 2 | Maternal ombilic – foetal ombilic |
| 2 | AS – foetal ombilic |
| 2 | Empty – fetus |
| 1 | Empty – foetal ombilic |
| 1 | Maternal ombilic – Foetal head |
| 1 | Maternal spine – foetus |
| 1 | Maternal abdomen – foetal ombilic |
| 1 | Placenta – fetal head |

### Interview: placenta perception

#### Placenta function and origin

The placenta was not perceived as an organ by a large majority of mothers (67%): « *Yes, it’s an organ, it allows him to do things that he’s not yet able to do, like breathing* » (E59); « *I think that the placenta is an organ, at least it has the essential functions* » (E38).

More than half of the mothers (53%) claimed that the placenta belongs to mother for a part and that it also belonged to the fetus for the other part. The two parts of the placenta constituted a maternal-fetal unit: « *I think it’s up to both of us, since if there is no pregnancy there is no placenta and without a placenta there is no baby* » (E6); « *For me, it is one of both* (…) *it’s the link between mother and baby, it’s something we share* » (E55).

During the interview, placenta concepts were explored, examples of which are presented below. Most mothers (77%) explained that the role of the placenta was to allow the growth of the fetus during pregnancy because it had a nutritional role: « (…) *this is how my baby takes nutrients from the diet so that he can develop*" (E36); « *It is the one who feeds the baby* (…) *through the cord*" (E12); « *It is the organ that will transmit all the nutritional exchanges between the baby and the mother* » (E13).

Although the placenta was presented as an organ that allows exchanges between mother and fetus (32%), it was more mentioned as a protection for the fetus (38%): « *It must have either a special contribution or a protective role* (…) *a role of defender whatever the context against the outside* (…) *an immune role, a barrier role* » (E58); « *It’s a bit like the filter between everything that comes from the outside and wants to go all the way to the little inside* » (E10);

« *It is used to make exchanges between mother and baby to carry to the child everything, he needs* » (E42); « *It’s an organ that is stuck against the uterine wall and is the link between mother and baby. It is connected to the umbilical cord* » (E54).

The placenta was implicated in the occurrence of a miscarriage at the beginning of pregnancy or bleeding during childbirth and its sequelae (20%): « *It can cause miscarriages* (…). *I had a placental hematoma and as a result it can cause bleeding, and you can have a miscarriage* » (E4); « *There must be no left in the uterus at all, otherwise it will cause hemorrhages* » (E12).

Eleven mothers (18%) said they did not know what the role of the placenta was during pregnancy and childbirth, and they did not provide an answer. Four mothers proposed menstruation or female hormones to explain origins and development the placenta.

#### Placenta location

Half of the mothers did not know where the placenta came from or how it developed. The origin of gametes was the most advanced explanation (18%): « *It is created with the egg system and at the very moment of fertilization, when everything is linked, there is surely a part of it that must be reserved for the creation of the placenta and that will come to attach itself to the uterus* » (E53); « *When fertilization occurs, spermatozoa meet an egg, then they migrate into the uterus and at that moment they form the placenta* » (E55).

Equal parts of mothers thought that the placenta came either from the uterus (10%) or from the embryo (10%), while 8% invoked supernatural explanations: « *It may be the endometrium that is lining and forming the placenta* » (E12); « *It is formed at the same time as the baby, from the fetus, everything that can be produced, by the secretions of the baby* » (E45); « *I take it as something very, very magical and full of things that fall into place suddenly and happen* (…) *there are plenty of things that happen that cannot be explained* » (E26).

Six mothers, in their comments, had negative perception of the placenta. They associated it with a "weird" and "disgusting" thing. The implantation of the cord at the umbilicus, from the fetus to the placenta, was known by 63% of mothers, but 23% also reported a cord implantation connected from the mother to the fetus. In equal measure, the section of the cord was seen as separating the mother from the newborn (47%) and the latter from the placenta (48%).

#### Placenta description and visualization after birth

The placenta was described as a fleshy material assimilated to a piece of meat (35%) with a bloody appearance (32%) and 22% of mothers presented it as a piece of liver (23%): « *I find that it looks like a kind of cutlet, a bloody piece of meat* » (E26); « *I was thinking of a piece of calf’s liver, it made me think of this* » (E47).

Very largely, mothers did not see the placenta after the birth of their children, whether in the situation of a normal birth (79%) or during a caesarean section (89%). According to 88% of mothers, the professional who performed the birth does not show or prose to see the placenta to the parents after the birth and no explanation is given to them.

Four mothers asked to see their placentas and three professionals took the step of showing mothers their placentas, especially when there is a medical reason that justifies it: « *They showed it to me because as there had been a hematoma, the gynecologist who gave birth to me showed it to me so that I could visualize what had happened in the last month* » (E53).

More than half of mothers (53%) would appreciate to look at the placenta (59%), out of the curiosity on their part and to understand what a placenta is (50%):« *Just to get an idea because we’re not pregnant every day, we’re not going to see him again and then it’s part of us* » (E13); « *To see what’s in the belly and what it looks like because I hear about it and I see it at the feed, but I’ve never seen it in real life, live, at least I know what I’ve carried in my belly* » (E15); « *I would have been interested to see it since it’s something that is quite obscured while it’s important even physically* » (E26).

To the proposal of seeing their placentas after childbirth, 40% of mothers would refuse it. At this point, they preferred to give priority to seeing and being with their newborns (37%) than to looked at the placenta, which they considered visually unattractive, even repulsive (29%) and uninteresting (25%). « *Not to see him especially, my daughter who came* (…) *I especially wanted to see my daughter in my arms more than anything else* » (E39); « *It would have disconnected me from the side of welcoming life, a baby* (…)*. The baby was there, we had finished everything that was technical, in quotation marks, because it’s often technical, there are doctors, things, we’re in the operating room, I don’t know, but bringing back a biological side is no longer too maternal, not too many bears. I needed this side to be a bit of a bear, more child-oriented, to ignore the medical side in fact* » (E57); « *I would have preferred it to be before the birth to explain to myself, at least before, what it was for. After childbirth, seeing him and understanding him is no longer very useful* » (E30).

However, they admit that 29% of them would be interested in seeing the placenta at a time further away from the birth of the child.

#### Placenta fate after birth : donation to research

The mothers talked about habits and customs around the placenta. Placentophagy in humans was the best known at 40%, followed by animal placentophagy (18%), the manufacture of therapeutics (15%), burial (12%) or preservation (7%) of the placenta. A good proportion of mothers (40%) had no knowledge or belief about placenta. In isolation, one mother evoked a family superstition related to infertility if the mother looked at her placenta and another mother talked about the practice of the lotus baby.

Ninety five percent of mothers had no personal or family aspirations related to the placenta that would require it to be retrieved after childbirth. Three mothers: a Senegalese, a Comorian and a French woman from Mayotte, mentioned that they might have wanted to recover the placenta to bury it and if they had wanted to follow the tradition or custom of the country.

More than half of mothers (53%) were unaware of the fate of their placenta after delivery and 90% would like to know what is really going to happen to them so they don’t stick to assumptions. Four mothers found no interest in knowing the fate of the placenta. Mothers who expressed a hypothesis were divided between donation for medical research (15%) and incineration with biomedical waste (30%).

The donation of placenta to scientific and medical research was mainly favored by mothers (93%). Sixteen percent had donated this organ in a humanistic way: *« It’s like donating your organs, it can save lives* (…) *to improve knowledge about the placenta for pregnancies that went wrong »* (E19) ; *« Yes* (…) *in the same way that one can donate blood or something else »* (E45) ; *« Yes, I am in favor of donation (…) for me, giving one’s body or a part of one’s body to science is essential for progress and for knowledge»* (E58).

Two mothers had mentioned the lack of information during the prenatal period and asked for information if a placental donation was considered. Four mothers had ethical considerations about donation: *« No, I’m not going to give it away if it’s not shown to me and if it’s not explained to me, it’s an organ that belongs to me »* (E25); *« There are sometimes research that goes a little too far for my taste, it would pose an ethical problem for me »* (E2); *« I don’t agree too much on too much science on the human body not to go towards robotism, as long as it’s not to harm »* (E34).

A request for placenta donation for industrial purposes was mostly refused by mothers (56%) for three reasons: the marketing of the donation (40%), the lack of transparency of industrial companies (20%) and 16% of them did not conceive of the use of the placenta in the manufacture of cosmetic products: *« If it’s trade for trade* (…) *it wouldn’t interest me, I really need to be explained precisely why it should be done »* (E57); *« No* (…) *it’s as if I were asked to give my blood for commercial purposes, I would say no, it’s to help others they don’t make money in my account »* (E36); *« This thing is pretty disgusting, and I think there are already enough weird products in cosmetics »* (E53).

### False beliefs and a significant lack of knowledge

The lack of knowledge about the placenta in our population was proven. The vast majority (67%) of mothers did not know what a placenta was. These mothers did not perceive the placenta as a transient organ whose expression "transient" allowed them to justify that the placenta could not be an organ:*« The placenta is only used for a given period, whereas an organ is available for life »* (E18); *« I wouldn’t say it’s an organ because at the end of pregnancy it comes out, maybe more of a tool than an organ because it’s not there all the time »* (E45).

Only a third of mothers known the origin of the placenta or could summarily explain how it developed: *« It is from a few cells, it is the first cell that is fertilized by the spermatozoon that will give a part for the placenta and [part] for the baby »* (E10);*« When fertilization occurs, spermatozoa meet an egg, then they migrate into the uterus and at that time they form the placenta »* (E55).

The placenta was completely obscured from their mental representations as shown by the graphic exercise. Most mothers, the placenta was clearly absent from their psychic schemas, since 77% of them did not represent it in graphic productions. It was the uterus that was mainly represented, which could refer to the psychoanalytic notion of the psychic envelope contained during pregnancy [14], which they associated with a representation of the umbilical cord that is connected to it. The actual implantation of the cord at the placenta, which was mostly absent, cannot therefore be drawn. The implantation of the cord could then be conceived as the fiction of the physical connection between the mother and her child where each end of the cord was then connected from the fetus to a part of its mother’s body: the maternal back or abdomen most often. This attachment between the maternal body and her fetus could symbolize the bond that the cord represents between mother and child during the prenatal period [14] since 47% of mothers claimed that the cord section separates the mother from the newborn: *« We cut the two parts that are connected to the cord. The mother’s body and the baby’s body, therefore the mother and the baby »* (E4); *« We separate the bond between mother and baby, we cut off all his [*baby’s] relationship *with me »* (E32).

Less than half of the mothers gave a reasoned explanation for the section of the cord separating the newborn from the placenta. However, when the placenta was represented in graphic productions, the cord almost always connected the placenta and the fetus (13 times out of 14). The collective imagination of a child’s loss by his mother at the very moment of his birth [15] remained anchored in the representations of almost half of our population. This emotional and symbolic dimension of separation referred to the idea of a fusional relationship between the mother and her fetus, for which 53% of mothers imagined that half of the placenta belongs to them and only the other half belongs to the fetus, which probably hindered the mechanisms of representation in biological and anatomical knowledge. Medicine referred to the term "feto-placental unit" as the anatomical unit from which the distinction between the embryo and the trophoblast will be established at an early age, [14] whereas more than half of mothers attributed a part of this anatomical unit to themselves in their accounts.

Some mothers (38%) perceived the placenta as a filter that would provide the fetus with immunological "protection" against bacteria and microbes. However, the expression "placental barrier", widely used in the popular imagination but also among professionals, was largely outdated. The scientific community had described the extent to which the placenta can cross gases, pathogens, drugs, toxins … but also maternal and fetal cells if its integrity was impaired [2,8]. The authors stressed that while there was probably also a form of barrier which protects, unfortunately in a very incomplete way, against certain infectious agents, it was now necessary to abandon this image and invent a new one, because this placental barrier was imperatively permeable.

The poverty of graphic productions reflected the general lack of knowledge of mothers. The graphic score was below average in 80% of mothers. The graphical representations were the elements that they understand [11] and that we had analyzed with the interviews. Thirty-four of these 48 mothers had only drawn their fetus or the fetus in the womb (**Figure 1**). The other mothers (14/48) added as elements to their drawings mainly the umbilical cord without attachment from the fetus or connected it in a whimsical way to themselves (**Figure 1**). When the placenta was drawn, it was most often correctly connected to the fetus (**Figure 1**).

**Figure 1.**
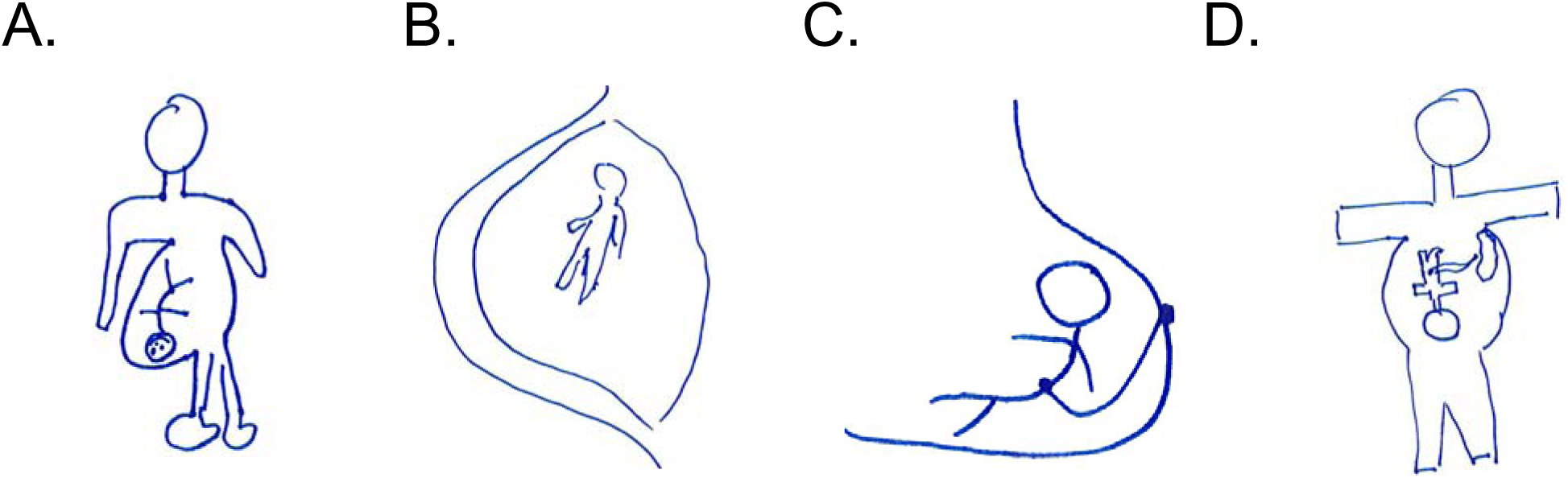
Drawing production. Graphic production of (**A**) the fetus (E24), (**B**) of the fetus in the womb (E21), (**C**) of the mother and the fetus connected by the umbilical cord (E35) and (**D**) of the fetus connected to the placenta by the umbilical cord (E14).

This lack of knowledge could explain the simple description of the placenta by mothers, who described it mostly as a piece of bloody "meat" or "liver". Without anatomical knowledge and explanation of the different physiological functions of the placenta, it seemed difficult to give a more elaborate description of the deliverance than that of its repellent aspect for some mothers once she had expelled it at delivery: *« It’s a bit of a weird aspect »* (E4); *« It’s a bit of a thaw but it’s not supposed to be pretty »* (E38); *« That thing was in me. It’s a bit weird this thing, I was impressed in fact (…) I hallucinated what it represents, it’s charming* [ironic tone]. *It’s a really disgusting thing »* (E53).

### Some real knowledge but not very constructed

The exchange of nutrients and products of fetal metabolism through the placenta was essential for fetal growth. The general mechanisms by which the exchange takes place were well-described in the literature and known by a large majority of mothers, who attributed a nutrient role to the placenta where the exchanges that serve the development of the fetus will be organized. Much of the physiology underlying these exchange mechanisms was still not known in detail by scientists. For the authors, a lack of knowledge was an obstacle to understanding the pathophysiology of the dysfunctional placenta, for example in the case of growth retardation [16]. However, 10% of the mothers interviewed discern, without explaining it, a causal link: *« If it doesn’t work well, it has effects, especially for the baby, it causes growth retardation »* (E4); *« It’s very bad if the food doesn’t get to the baby, it can do a lot of damage to the size of the little one, to the development of its size »* (E43).

A proportion of mothers (20%) were aware that bleeding can occur if the placenta had not been completely evacuated from their bodies during delivery or during its aftermath and cause hemorrhage.

Only slightly more than half of the mothers had any knowledge about the placenta other than bio-medical. The practice of placentophagy was the most reported among these mothers because it had been publicized across the Atlantic by reality TV stars or in fiction. This practice was also associated with the imagination of primitive tribes: « *I know there are some who want to eat it like Kim Kardashians. I know that there are some who eat it apparently it has great protein virtues* » (E51); « *If* (…) *in movies where some people eat their placentas. If in horror movies in the United States* » (E57); « *In some countries there are those who consume it raw, in underdeveloped countries, in tribes in Africa* » (E48); « *I imagine that there are primitive societies that eat it, if it contributes to immunity why not. They eat it to recover the strength in quotes* » (E26).

Placentophagy was generally prohibited in human societies, whereas it was normal in animals but less often mentioned by mothers. Since the 1970’s, mainly in the USA, a return of this practice had been noted in spiritual movements that advocate natural childbirth at home [15,17]. The practice was controversial, and no benefit was documented to recommend it [18]. In some French territories, a request from mothers or families consisted of being able to retrieve the placenta after childbirth to be able to bury it. In mainland France, this demand remained a minority and most often comes from Polynesian, Comorian or African families, whose practice was disappearing with the acculturation of generations [6]. In our study, no mother asked to retrieve her placenta. The perpetuation of a custom through the rites of burial or burial linked to a country of origin, evoked by three mothers, seems to rival French health protocols only in the territory of origin. There was a shift in the individual or family meaning of a practice linked to the cultural and post-natal management of the placenta towards indigenecity and the nationalist assertion of identity claims [7]. In France, the question of the status and fate of the placenta was not up for debate. More than half of mothers did not know what happens to it after birth and barely 30% know that it was considered "hospital waste" and was cremated. Nine mothers think that it was given to medical research without really "knowing why". This was a possibility requiring a protocol for researchers. The status of "transitional organ" of the placenta did not allow, under current legislation, placental donation to be included in the customary practice of organ donation. Most mothers were largely in favor of medical research and placental donation to science, regardless of their origins and personal convictions. Legislative adaptation was necessary and raises ethical questions [19]. These questions arise because we were now witnessing a "renaissance of the placental ritual" nourished by the belief in the present or future virtues of cord blood stem cells, with which Science allowed families to envisage regenerative medicine [20]. Donation of the placenta, as with the other organs in their collections, was already assimilated by some mothers (9/60) as an "ordinary donation".

### Management of the placenta by hospital professionals

We did not find any studies that investigated or described caregivers’ practices for managing the placenta following delivery. Hospital health protocols were applied by the professionals who delivered the birth to the mother. After delivery and having ensured the integrity of the dispensing, they evacuated the placenta, considered as a potentially infectious surgical waste, in a waste disposal container to be then treated by incineration. The evacuation of the placenta was mostly done without the mothers’ knowledge. In 53 interviews out of 60 deliveries, the professionals neither showed nor offered to see their placentas to the mothers. Only four mothers asked the caregivers to be able to see their placenta following delivery. A recent study looked at the experience of mothers during delivery [21]. Mothers reported that they had not received any information on the management of the third stage of labour and health professionals did not provide information until the delivery was managed. This study reflected the usual practices of caregivers who did not involve mothers in the decisions to be made, whereas the French code of ethics (Art. R.4127 CSP) required all midwives to inform their patients and to respect their wishes as far as possible. In our study, only three professionals took the initiative to show mothers their placentas, because this was in two cases the explanation of the situation: *« We were shown it right away because afterwards we see the baby because we had to go and get the cord, the ball with all the equipment in it. But then we are not given it »* (E3); *« They showed it to me because there was the hematoma, the gynecologist who gave birth to me showed it to me so that I could visualize what had happened in the last month »* (E53).

## Discussion/Conclusion

There were limitations to our study. Qualitative methods of data collection encouraged researchers to remain cautious and not to draw general conclusions. However, beyond 40 interviews, it was reasonable to think that a saturation of data for a population can be envisaged in its study context [12]. With 60 interviews, we suggested that our results generally represent the responses of the women who had given birth in the Marseille maternity hospitals in which we interviewed them. Our study offered results that had been little studied in medicine and midwifery. Experts and professionals allowed themselves to say that it was enough to: "*Ask a pregnant woman, she is unaware of the existence of this organ* [of the placenta] *in most cases*" [2,15]. This statement was not based on any study describing knowledge of the fetal appendages or the placenta in mothers. The theme also suffered from a deficit of thought and representations in the human sciences [14] that it seemed appropriate to study.

Even if more than half of mothers would appreciate seeing their placentas and obtaining information about them from the caregiver who performed the delivery, it did not seem reasonable to us at present to require this from professionals. For several years now, professional organizations had been calling for a revision of the 1998 decrees that regulate the number of professionals present in the perinatal sectors and denounced a glaring lack of caregivers with repercussions for the safety of users [22,23]. This desire of mothers to receive information about their placentas could find its place upstream during pregnancy without becoming a systematic approach for caregivers since 40% of the mothers we interviewed will decline the offer to see the delivered. The study by Reed et al. confirmed that caregivers most often fail to did so due to lack of time [21], knowing that in our study 38% of mothers preferred to welcome and saw their newborns after delivery than to look at the placenta, which they found uninteresting at 25%. Institutional and health guidelines on the management of the placenta should be discussed with users. They would benefit from being presented before delivery. Since 2019, a preventive prenatal check-up had been added as part of the pregnancy follow-up. It allowed self-employed midwives to carry out preventive acts as part of a 45-minutes interview reimbursed at 100% by social security. This interview should preferably be carried out at the beginning of pregnancy to meet its objectives, particularly about hygienic and dietary rules. It made it possible to discuss measures to prevent toxoplasmosis in HIV-negative mothers. During BPP sessions, a time should address the management of delivery and management of the placenta with the professional present at the birth. If mothers so wish, they should be encouraged to be proactive in their requests to be able to access their placentas to look at them and obtain information. This approach seemed reasonable to us because it involved mothers more in making decisions with caregivers in the organization of their deliveries, contributing to making it a successful positive experience [24].

This study provided new data on the knowledge and representations on the placenta from women who have given birth during their pregnancies and following delivery. Most mothers had a major lack of knowledge and misrepresentations about what a placenta was and its different functions. These shortcomings make it difficult for mothers to produce this organ that they considered essential for the development of their children but that they were ready to give to science "if it can help" once the birth was over. In France, mothers did not seem to have any complaints for the placenta following birth, but cultural practices exist and could be heard. An increase in the representation of the placenta by mothers appeared necessary for better management of delivery and practices around the placenta with them, especially for mothers who wished to give birth at home or in a birthing center. Studies showed that when mothers choose to experience physiological or "natural" childbirth in these places, they showed a renewed interest in the placenta and its management [17–19,21].

## Data Availability

All data produced in the present study are available upon reasonable request to the authors

## Author Contributions

### Funding

No funding to declare.

### Data Availability Statement

The authors confirm that the data supporting the findings of this study are available within the article and its supplementary materials.

